# Association of VEGF and KDR polymorphisms with the development of schizophrenia

**DOI:** 10.1101/2021.08.06.21261566

**Authors:** Hana Saoud, Youssef Aflouk, Amira Ben Afia, Lotfi Gaha, Besma Bel Hadj Jrad

## Abstract

**Aim:** Several approaches indicate different blood flow disturbances in schizophrenia (Scz). Vascular endothelial growth factor (VEGF) is widely recognized as one of the key molecules implicated in the angiogenesis process through mainly its receptor KDR. The current work was designed to investigate the potential association between three polymorphisms (rs699947; rs833061 and rs3025039) in VEGF gene and two SNPs (rs2305948 and rs1870377) within KDR gene and predisposition to Scz among the Tunisian population.

**Methods:** We carried-out a case-control study composed of 200 schizophrenic patients and 200 healthy subjects using RFLP-PCR.

**Results:** Of all analyzed polymorphisms, only rs3025039, rs833061 and rs1870377 showed a significant risk for Scz. Following the stratified analysis, rs833061 was more prevalent among undifferentiated form. Yet, rs1870377 was especially correlated with paranoid subtype. We found also that rs699947 and rs833061 had an impact on patients’ symptomatology. Haplotype analysis unveiled a strong LD between rs833061 and rs3025039 only for undifferentiated patients. Moreover, the -2578/-460/+936 CTT haplotype, with only one mutated allele +936T, conferred a high risk to Scz and, in particular, to undifferentiated and paranoid forms. Among the last-mentioned subgroup, we noticed another overrepresented haplotype (ATT). Furthermore, the +1192/+1719 GT haplotype carrying the minor allele +1719T displayed increased frequencies in schizophrenics as well as in paranoid patients.

**Conclusion:** Our results show that all SNPs associated with the development or the severity of schizophrenia, were subsequently correlated with a decrease in the VEGF levels or influence VEGFR-2 binding affinity. Nevertheless, these data need to be strengthened by further independent analyses.

## 1) Introduction

Schizophrenia (Scz) is a complex neuropsychiatric disease characterized by a variable symptomatology with several clinical forms and progressive modes. The etiology remains poorly elucidated despite extensive research and abnormalities involved in the development of this disabling disorder include a variety of complex pathways. Accumulating data have indicated over-expressed genes implicated in vascular function, vasoregulation, shear stress, cerebral ischemia, neurodevelopment and post-ischemic repair among genes that might predispose the risk for the development of Scz^1^. Moreover, neuroimaging approaches have identified dilated lateral ventricles and smaller tissue volumes in various brain regions which can possibly be explained by hypoperfusion or diminished blood circulation observed in schizophrenic subjects^2,3^. On the other hand, cognitive deficits were ameliorated after injection of recombinant human erythropoietin, known to stimulate the process of angiogenesis^4,5^. Aside from being a potent angiogenic factor, vascular endothelial growth factor (VEGF) plays a relevant role in the central nervous system^6^ and is secreted not only by endothelial cells, but also by astrocytes and neurons of any degree of maturity^7^. It acts via three specific tyrosine kinase receptors: VEGFR1, VEGFR2 and VEGFR3^8^. VEGFR2, alternatively designated as KDR (*kinase insert domain receptor*), is recognized as the major signal transducer and plays a main role in mitogenic, angiogenic and permeability-enhancing effects of VEGF^9,10^. As well, this receptor contributes to the neurotrophic actions of VEGF, which promotes axonal outgrowth, enhances cell survival and proliferation^11^. *Post-mortem* findings unveiled lower VEGF mRNA levels in the dorsolateral prefrontal cortex of patients with Scz^12^. Indeed, genetic variation in VEGF could impact human hippocampal morphology^13^. This brain area has long been regarded a key region in the pathophysiology of this psychosis^2,14^ and physiological abnormalities of the hippocampus^15,16^ are well established in individuals suffering from Scz. Additionally, the inflammatory mediators may alter the integrity of the blood-brain barrier through reactivation of VEGF signaling^17^ and several pro-inflammatory cytokines, which are suspected to play a role in the vulnerability to Scz, might impact VEGF levels^18^.

The VEGF gene is mapped to chromosome 6 (6p21.3)^19^ and includes several polymorphisms that were of particular interest to us: -2578C/A (rs699947); -460T/C (rs833061); +936C/T (rs3025039) that may influence VEGFA expression. Besides, the gene encoding KDR is located on chromosome 4 (4q11-q12)^20^. There is conclusive data demonstrating that functional genetic variants (+1192G/A (rs2305948); +1719A/T (rs1870377)) within KDR gene may affect biological function of this receptor. Therefore, the aim of the present study was to evaluate the relationship between the above-mentioned polymorphisms and Scz, and to assess the possible relationships between genotypes and clinical characteristics of this psychotic disorder.

## 2) Material and Methods

### 2.1. Study Participants

A group of 200 patients with the diagnosis of Scz (17.8 % female; 82.2 % male; Sex ratio: 4.6; mean age: 38.21 ± 10.83 years) were enrolled in the current work. The diagnosis of the pathology was based on the criteria mentioned in the DSM-IV axis I^21^ with an accuracy of sub-clinical type. 48.9% of cases met the criteria for undifferentiated, 31.5% for paranoid and 18.3% for disorganized forms of Scz. Our study omitted patients with acute illness (allergic, autoimmune or malignant disorders) or other comorbidities such as epilepsy, bipolar and depression. Showing continuous signs of the disturbance for minimum six months with active symptoms (hallucinations, delusions, etc.) for at least one month is an essential condition to be admitted as true schizophrenic. Information about the age of onset, which corresponds to the appearance of the first psychotic symptoms, and total scores for positive, negative and general syndromes were documented.

Patients were recruited from the Psychiatry Department of Monastir Hospital, Tunisia whereas healthy volunteers were selected from the regional blood transfusion center of the same University Hospital. The control group included 200 subjects with a mean age of 33.76 ± 10.79 years and a sex ratio of 3.6 (21.5% females and 78.2% males). The main exclusion criteria were as follows: current neurological or chronic physical problems, family history of psychiatric disease and substance abuse.

The study was authorized by the research in life sciences and health (CER-SVS) ethical committee of the Higher Institute of Biotechnology of Monastir. Informed consent was obtained from each participant or a family member prior to blood sampling.

### 2.2. Genomic DNA analysis

Blood samples from all participants were collected in an EDTA-containing tube and the genomic DNA was extracted using the salting-out technique^22^. Regions covering targeted polymorphisms were amplified by the RFLP-PCR. Based on prior reports, PCR primers, restriction enzymes, and product sizes were designed^23,24,25^ and details are listed in Table 1.

**Table 1:**
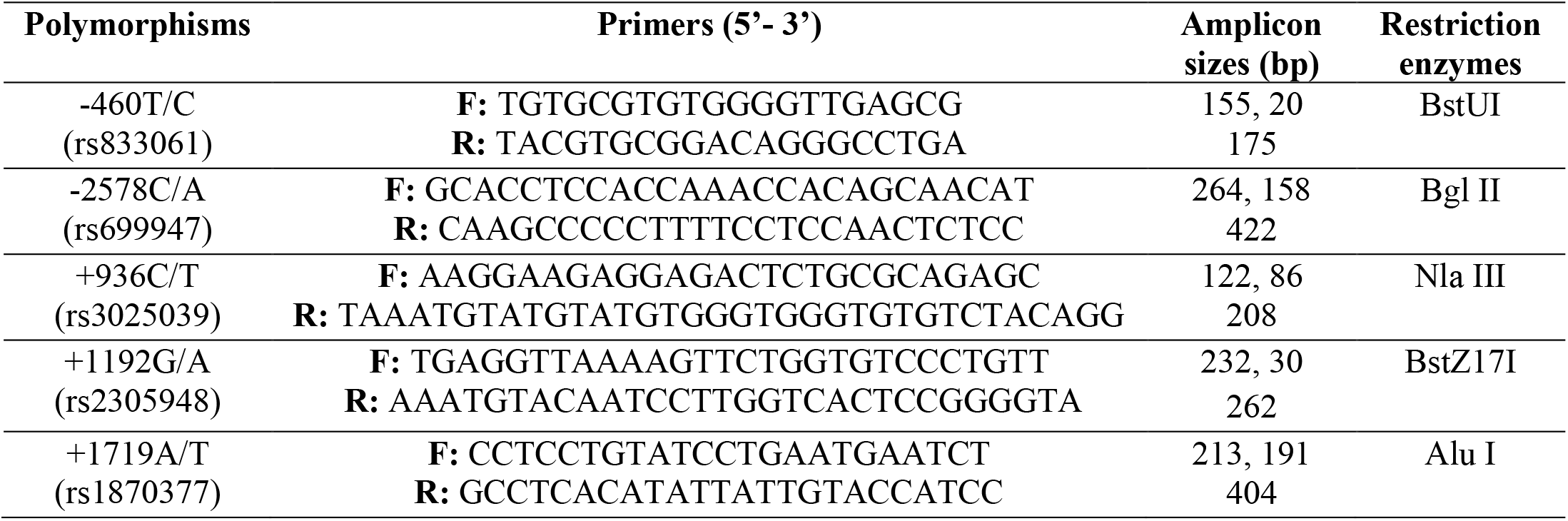
Primers of VEGF and KDR gene polymorphisms for PCR amplification

### 2.3. Statistical Analysis

The Hardy-Weinberg equilibrium (HWE) was estimated for the studied SNPs in both groups using the Chi-square test. For statistical analysis, SPSS software (version 23, Armonk, NY, USA) and SNPStats online software (https://www.snpstats.net/start.htm) was performed. The strength of association between alleles or genotypes of control and case samples was evaluated with the odds ratio (OR) presented with 95% confidence intervals (CI) whenever χ2 test was significant. Statistical significance was assigned at p < 0.05.

The best model of inheritance was determined according to the lowest Akaike Information Criterion (AIC) value^26^ (Table 2). Additionally, students’ t-test was designed to compare symptom severity scale in schizophrenic patients according to VEGF genotypes before and after treatment. P-value adjustment (PA) (for the effects of age and gender) and haplotype reconstruction were carried by SNPStats. Linkage disequilibrium (LD) expressed in terms of the D′ and r^2^ coefficients was estimated between paired SNPs by the same online software.

**Table 2:** Different frequencies of VEGF and KDR polymorphisms between patients and controls according to the different models of inheritance

| Inheritance Models | Genotypes | Controls n (%) | Patients n (%) | OR (95% CI) | P-value | AIC | P<sub>A</sub> |
| --- | --- | --- | --- | --- | --- | --- | --- |
| **VEGF-A -2578 C/A (rs699947)** |  |  |  |  |  |  |  |
| Codominant | CC | 72 (36%) | 72 (36%) |  |  |  |  |
|  | CA | 105 (52.5%) | 97 (48.5%) | 0.92 (0.6 - 1.4) |  |  |  |
|  | AA | 23 (11.5%) | 31 (15.5%) | 1.35 (0.7 - 2.5) | 0.47 | 559 | 0.62 |
| Dominant CC vs CA+AA |  |  |  | 1 (0.6 - 1.5) | 1 | 558.5 | 0.65 |
| Recessive CC+CA vs AA |  |  |  | 1.41 (0.7-2.5) | 0.24 | **557.1** | 0.49 |
| Alleles | C | 249 (62.25%) | 241 (60.25%) |  |  |  |  |
|  | A | 151 (37.75%) | 159 (39.75%) | 1.08 (0.8 - 1.4) | 0.56 |  | 0.50 |
| **VEGF-A -460 T/C (rs833061)** |  |  |  |  |  |  |  |
| Codominant | TT | 137 (69.9%) | 119 (59.5%) |  |  |  |  |
|  | TC | 56 (28.6%) | 76 (38%) | 1.56 (1.02 - 2.3) |  |  |  |
|  | CC | 3 (1.5%) | 5 (2.5%) | 1.92 (0.45 - 8.2) | 0.09 | 550.2 | 0.13 |
| Dominant TT vs TC+CC |  |  |  | 1.58 (1.04 - 2.4) | 0.03* | **548.2** | 0.04* |
| Recessive TT+TC vs CC |  |  |  | 1.65 (0.39 - 7) | 0.49 | 552.5 | 0.49 |
| Alleles | T | 330 (84.2%) | 314 (78.5%) |  |  |  |  |
|  | C | 62 (15.8%) | 86 (21.5%) | 1.45 (1 - 2.09) | 0.04* |  | 0.04* |
| **VEGF-A +936 C/T (rs3025039)** |  |  |  |  |  |  |  |
| Codominant | CC | 157 (78.5%) | 122 (61%) |  |  |  |  |
|  | CT | 41 (20.5%) | 69 (34.5%) | 2.17 (1.3 - 3.4) |  |  |  |
|  | TT | 2 (1%) | 9 (4.5%) | 5.79 (1.2 - 27.2) | 0.0003* | 544.1 | 0.0013* |
| Dominant CC vs CT+TT |  |  |  | 2.33 (1.5 - 3.6) | 0.0001* | **543.8** | 0.0005* |
| Recessive CC+CT vs TT |  |  |  | 4.66 (1 - 21.8) | 0.026* | 553.6 | 0.062 |
| Alleles | C | 355 (88.75%) | 313 (78.25%) |  |  |  |  |
|  | T | 45 (11.25%) | 87 (21.75%) | 2.19 (1.4 - 3.2) | 0.00006* |  | 0.0002* |
| **KDR +1192 G/A (rs2305948)** |  |  |  |  |  |  |  |
| Codominant | GG | 150 (75%) | 149 (74.5%) |  |  |  |  |
|  | GA | 43 (21.5%) | 47 (23.5%) | 1.1 (0.6 - 1.7) |  |  |  |
|  | AA | 7 (3.5%) | 4 (2%) | 0.58 (0.1 - 2) | 0.6 | 559.5 | 0.64 |
| Dominant GG vs GA+AA |  |  |  | 1.03 (0.6 - 1.6) | 0.91 | 558.5 | 0.61 |
| Recessive GG+GA vs AA |  |  |  | 0.56 (0.1 - 1.9) | 0.36 | **557.7** | 0.54 |
| Alleles | G | 343 (85.75%) | 345 (86.25%) |  |  |  |  |
|  | A | 57 (14.25%) | 55 (13.75%) | 0.95 (0.6 - 1.4) | 0.83 |  | 0.84 |
| **KDR +1719 A/T (rs1870377)** |  |  |  |  |  |  |  |
| Codominant | AA | 114 (57%) | 93 (46.5%) |  |  |  |  |
|  | AT | 74 (37%) | 81 (40.5%) | 1.3 (0.8 - 2) |  |  |  |
|  | TT | 12 (6%) | 26 (13%) | 2.6 (1.2 - 5.5) | 0.021* | 552.8 | 0.025* |
| Dominant AA vs AT+TT |  |  |  | 1.5 (1 - 2.2) | 0.035* | 554.1 | 0.023* |
| Recessive AA+AT vs TT |  |  |  | 2.3 (1.1 - 4.7) | 0.016* | **552.7** | 0.03* |
| Alleles | A | 302 (75.5%) | 267 (66.75%) |  |  |  |  |
|  | T | 98 (24.5%) | 133 (33.25%) | 1.5 (1.1 - 2) | 0.006* |  | 0.005* |

**OR:** odds ratio; **CI:** confidence interval; \***P**< 0.05; **P<sub>A</sub>:** Adjusted P-value for age & sex **AIC** (Akaike information criterion) provides a means for model selection.

## 3) Results

The distribution of VEGF and KDR genotypes were in keeping with those predicted by the Hardy-Weinberg equilibrium conditions in both healthy controls and patients (data not shown). The genotypic and allelic frequencies of investigated SNPs are depicted in table 2. Statistical analysis unveiled no notable differences between the promoter allele -2578A and predisposition to SCZ (PA=0.5). As for the second SNP-460T/C, we note that the minor C allele and the combined (TC+CC) genotype are more prevalent among cases (PA=0.04 and PA=0.04, respectively). With regard to +936C/T substitution, a strong association was seen for TT genotype under the three inheritance models (table 2). In terms of allele frequency, an increased risk of schizophrenia was perceived (PA=0.0002).

Concerning the +1192G/A variant of the KDR gene, the obtained results evoked close values for both groups, indicating no significant association between the genotyped KDR mutation and SCZ. Instead, the lower prevalence of the mutated TT genotype (whatever the genetic mode of inheritance) as well as the minor T allele in healthy subjects suggests that the +1719A/T polymorphism constitutes a risk for the development of this pathology.

These positive correlations, reinforced by adjusted analysis for age and gender, remain highly significant with the lowest Akaike information criterion (AIC) conforming to the dominant model of inheritance for the -460T/C, +936C/T polymorphisms and to the recessive one for the +1719A/T polymorphism (Table 2).

According to data analysis of clinical subtypes, no significant association was retrieved for polymorphisms at positions -2578C/A, -460T/C and +1192G/A. However, a higher distribution of +936C/T was found among undifferentiated and paranoid forms considering the dominant inheritance. As well, the +1719A/T substitution was correlated with paranoid and disorganized schizophrenia but not with the undifferentiated type of the disease conforming to the recessive mode of inheritance (Table 3).

**Table 3:**
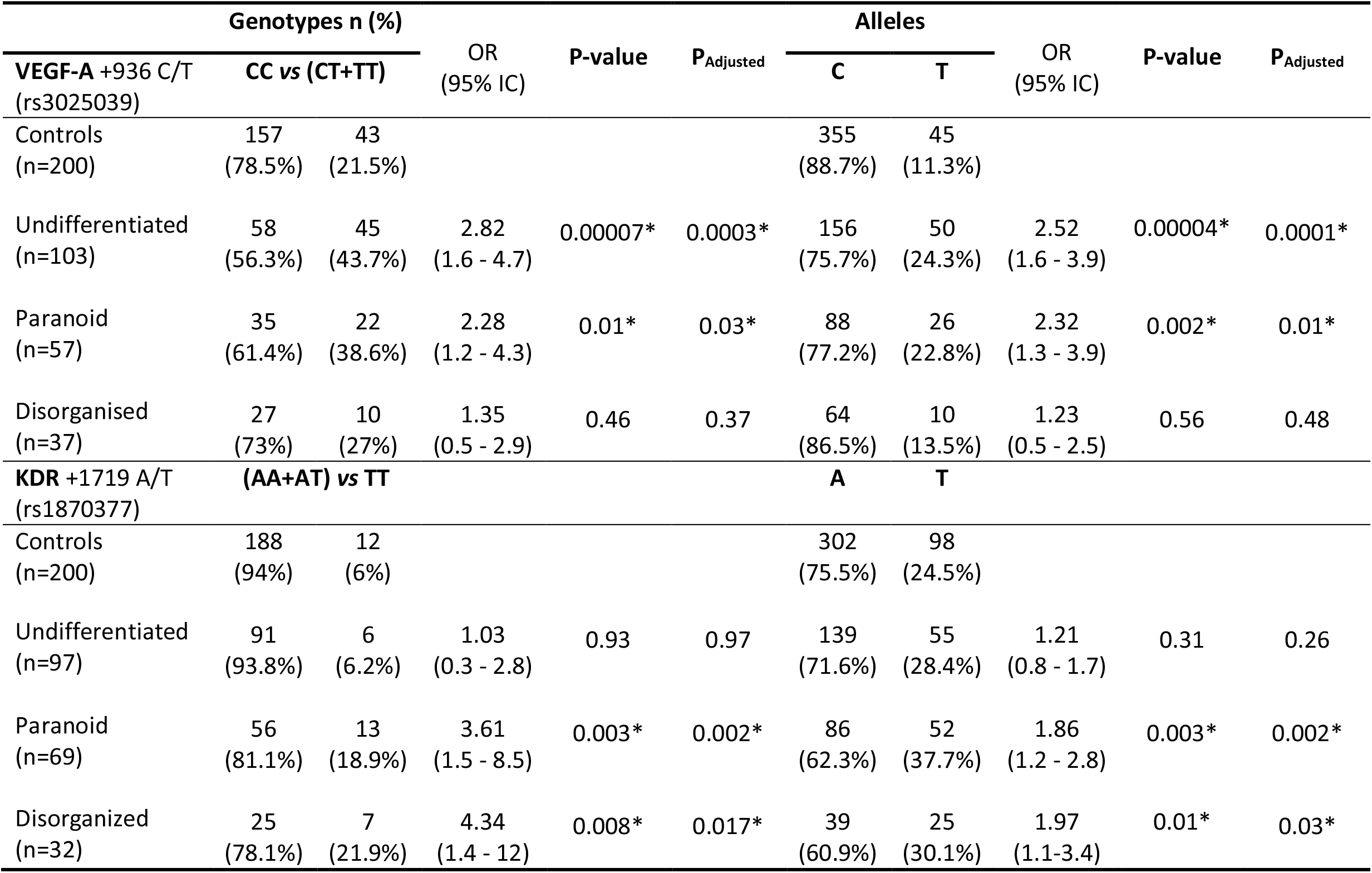
Distribution of genotype and allele frequencies of rs3025039 & rs1870377 in control subjects and patients with different subtypes of Scz

When we performed a sex-stratified analysis, our results indicated that only undifferentiated males harboring +936T allele and paranoid males carrying +1719T allele are more predisposed to SCZ as expressed by the appropriate model of inheritance of each polymorphism (***PA***=0.0003; ***PA***=0.005 respectively). Similar results have been described for females without reaching the statistical significance; one should take into account the relatively small size of the designed population. However, allele differences were not significant for both sexes in terms of the other analyzed sites (data not shown).

In order to find out if there is an impact of the -2578C/A polymorphism on patients’ symptomatology, we compared the severity of disease symptoms before and after treatment. As shown in table 4, cases with AA genotype have more intense symptomatic scores at admission and discharge with statistical significance for the SANS1 (p=0.004) and SANS2 (p=0.001) scales according to the recessive genetic model.

**Tableau 4:** Comparison of symptom severity scale in schizophrenic patients according to the recessive model of the -2578 C/A SNP before (1) and after (2) treatment (mean ± standard deviation)

|  | Genotypes |  |  |
| --- | --- | --- | --- |
| | CC+CA (n) | AA (n) | $p$ (CC+CA vs AA) |
| SAPS 1 | 32.4 ± 5.6 (125) | 33.5 ± 7.2 (25) | 0.4 |
| SANS 1 | 33.2 ± 6.3 (123) | 37.4 ± 6.1 (24) | 0.004* |
| BPRS 1 | 37.7 ± 3 (129) | 37.9 ± 3.9 (27) | 0.76 |
| SAPS 2 | 25 ± 7.1 (134) | 25.2 ± 8 (25) | 0.9 |
| SANS 2 | 26.3 ± 8.1 (132) | 32.4 ± 7.6 (23) | 0.001* |
| BPRS 2 | 33.2 ± 3.8 (139) | 33.5 ± 5.9 (27) | 0.78 |

**BPRS:** Brief Psychiatric Rating Scale; **SANS:** Scale of the Assessment of Negative Symptoms; **SAPS:** Scale of the Assessment of Positive Symptoms.

Similarly, heterozygous and homozygous carriers of VEGF +936C/T presented greater positive and negative symptoms score means. The difference is statistically significant after treatment as mentioned in the table 5. For the -460T/C, +1719A/T and +1192C/T SNPs we did not find any significant correlation with the scores of the psychometric parameters (BPRS, SANS and SAPS) whatever the genetic transmission mode.

**Tableau 5:** Comparison of symptom severity scale in schizophrenic patients according to the dominant model of the +936 C/TSNP before (1) and after (2) treatment (mean ± standard deviation).

|  | Genotypes |  |  |
| --- | --- | --- | --- |
|  | CC (n) | CT+TT (n) | *p* (CC vs CT+TT) |
| SAPS 1 | 32.1 ± 5.5 (90) | 33.1 ± 6.6 (61) | 0.3 |
| SANS 1 | 33.2 ± 6.9(86) | 34.6± 5.8(61) | 0.2 |
| BPRS 1 | 37.9 ± 3 (94) | 37.9 ± 3.3 (63) | 0.9 |
| SAPS 2 | 24.1 ± 7.3 (96) | 26.7 ± 7.5 (61) | 0.03* |
| SANS 2 | 25.5 ± 8.2 (92) | 29.1 ± 7.9 (60) | 0.008* |
| BPRS 2 | 33.3 ± 3.5 (101) | 33.8 ± 4.3 (63) | 0.4 |

We conducted haplotype analysis in order to estimate the combined effect of the VEGF and KDR polymorphisms. The -2578/-460/+936 CTT haplotype, with only one mutated allele +936T, conferred a high significant difference in patients (P=0.016). Furthermore, this genetic combination exerts much important predisposing effect in the undifferentiated and paranoid forms (P= 0.03; P=0.02 respectively). Among the last-mentioned subgroup, we noticed that -2578/-460/+936 ATT haplotype is overrepresented (P= 0.01). Moreover, the +1192/+1719 GT haplotype carrying the KDR +1719T displayed significant increased frequencies in schizophrenics as well as in paranoid patients (p=0.013; p=4e^-04^*respectively).

In connection with the previous results, haplotype distribution profile of the +936C/T and +1719A/T polymorphisms suggests that the observed predisposing effect may be driven by the VEGF+936T allele. However, among the paranoid subgroup the +936/+1719 TT haplotype carrying two mutated alleles conferred the highest significant difference as compared to the wild type combination. Estimated constructed haplotypes are shown in table 6.

**Table 6:** Haplotype frequencies of the rs 3025039 and rs 1870377 in healthy controls, patients with Scz and according to paranoid subtype

| KDR<br>haplotypes | Percent haplotype<br>frequencies |  | OR (95%CI) | P-value |
| --- | --- | --- | --- | --- |
|  | Controls | SCZ patients |  |  |
| +936/+1719 |  |  |  |  |
| C-A | 67.8 | 53.3 |  |  |
| C-T | 20.8 | 24.7 | 1.4 (0.9 - 2.1) | 0.06 |
| T-A | 7.7 | 13.5 | 2.2 (1.2 - 4) | 0.01* |
| T-T | 3.6 | 8.3 | 3 (1.2 - 7.5) | 0.01* |
| KDR<br>haplotypes | Controls | Paranoid<br>patients | OR (95%CI) | P-value |
| C-A | 67.8 | 50.76 |  |  |
| C-T | 20.8 | 25.74 | 1.5 (0.8 – 2.6) | 0.13 |
| T-A | 7.7 | 11.69 | 1.8 (0.8 - 4) | 0.16 |
| T-T | 3.6 | 11.81 | 4.9 (1.8 - 13.6) | 0.0021* |

The D’ and r values for controls and undifferentiated patients are stated in table 7. There was no evidence of LD between markers except for -460T/C and +936C/T + (p=0,005).

**Table 7:** Linkage disequilibrium between the three analyzed VEGF SNPs

| **SNP** | **D' (r)** |  | **p-value** |  |
| --- | --- | --- | --- | --- |
|  | **VEGF-460T/C** | **VEGF+936C/T** | **VEGF-460T/C** | **VEGF +936C/T** |
| VEGF -2578C/A | 0.48 (0.29) | 0.03 (-0.01) | 0 | 0.81 |
| VEGF -460T/C | - | 0.14 (0.13) | - | 0.005* |

**r:** Pearson's correlation coefficient; **D':** Standardized linkage disequilibrium coefficient

## 4) Discussion

Several micro and macrovascular disorders, involving peripheral and cerebral vascularization have recently been identified as having direct effects on the pathophysiology of Scz. Researchers have focused on exploring plasma levels of some angiogenic factors such as VEGF in neuropsychiatric conditions like depression or bipolar type I disorder^27,28^. In doing so, we chose to analyze functional polymorphisms of the VEGF gene and its KDR receptor to identify the nature of the plausible link between Scz and angiogenesis from a vascular point of view. The VEGF gene, highly polymorphic, is localized on chromosome 6 at locus 6p12. In the promoter region, a C/A substitution is described at position -2578. The -2578C allele has a binding site to the HIF-1 transcription factor which can be removed by the -2578A minor allele^29^. HIF-1 is required to modulate the transcription of the gene in response to hypoxia via interaction with the HRE (*Hypoxia Response Element*) located in the 5’UTR region^30^. Moreover, the -2578A allele is linked to the insertion of 18bp at position -2549 (rs144854329)^31^ which is absent in individuals harboring the wild C allele^32^. The homozygous CC genotype increases the production of this growth factor in the PBMC (*peripheral blood mononuclear cell*) supernatant in culture from healthy subjects^33^. Furthermore, the -2578C/A SNP is involved in the development of some neurodegenerative disorders, such as Amyotrophic Lateral Sclerosis and Alzheimer’s disease^34,35^, as well as the severity of atherosclerosis^36^ and psoriasis^37^. As regards to mood disorders, this mutation is correlated with treatment-resistant depression^38^. In our prospective study, the comparison of genotypic and allelic frequencies did not reveal any significant association (*p*>0.05) with the development of Scz in the Tunisian population. This result suggests that the predisposition to this psychosis is unrelated to the production of VEGF in hypoxic condition. On the other hand, carriers of AA genotype display important SANS scale at admission (*p*=0.004) and at discharge (*p*=0.001) of patients. It means that recessive A allele, which down-regulate VEGF levels in hypoxic situations, confers the ability to have severe negative symptoms. According to a report on brain function, this genetic variation causes an attenuation of white and gray matter and arterial blood volume in healthy individuals^39^. Another work dealing with psychosis, including Scz, found that -2578C/A mutation is not associated with the intensity of positive symptoms and cognitive abilities of patients. Yet, they did not elaborate on negative symptoms^40^.

Regarding -460T/C SNP, we noticed that the minor C allele along with the combined TC+CC genotype are modestly more frequent in patients with schizophrenia compared to controls. This substitution, situated in the promoter region, enhances VEGF gene expression *in vitro* using the human breast cancer cell line MCF7^41^. It is implicated in the development of proliferative diabetic retinopathy, acute respiratory distress syndrome and endometriosis^42,43^. As for cancer susceptibility, results are often contradictory^44,45^.

We also investigate, for the first time, the involvement of +936C/T polymorphism detected in the 3’UTR region that up regulates VEGF plasma concentrations among carriers of the +936T allele^46^. This mutation leads to the loss of a potential binding site for the transcription factor AP-4 (*activating enhancer binding protein 4*)^47^. In the current study, we showed a risk for the development of Scz with +936T allele and the TT genotype considering the three inheritance models. This predisposition remained valid for the paranoid form and more advanced in the undifferentiated subtype even after adjustment with covariates. This result is similar to that found for MCP-1 -362G/C among the same population^48^. When we compared the severity of disease symptoms, our findings demonstrate also greater symptomatic scores at admission and discharge with statistical significance for the SAPS2 and SANS2 scales according to the dominant genetic model. In order to evaluate the combined effect of the three targeted SNPs, we carried out a haplotypic analysis. The obtained results unveil that -2578/-460/+936 CTT haplotype, with only one risk allele (+936T), present a statistical difference in the whole schizophrenic population (*p*=0.016) and persisted significant for the undifferentiated and paranoid group. We noted also that -2578/-460/+936 ATT is highly expressed among the last-mentioned clinical form.

As far as we know, this is the first report of these variants being examined for correlation with Scz. In agreement with our results showing that alleles associated with decreased production of VEGF predispose to the development or severity of Scz, *post-mortem* studies have shown a decreased VEGF brain activity among schizophrenics. Withal, levels of its mRNA in the PFC region as well as expression of its KDR receptor are significantly reduced^12,49^. VEGF acts as a neurotrophic factor in neurons and appears to play a critical role in adult neurogenesis^50,51^. In fact, the proliferation of neural stem cells in the hippocampus (first stage of adult neurogenesis) seems significantly restricted in this psychotic disorder^52^.

Referring to several subsequent works, serum levels are elevated whereas brain VEGF expressions are declined in individuals suffering from Scz. These results, looking contradictory, could be explained by mechanisms involving internalization and degradation of the VEGF/KDR complex that would inhibit cerebral VEGF signaling pathways^49^. Thus, the correlation proposed by our work between the two SNPs, generating elevated VEGF levels, and susceptibility to Scz would be consistent.

This study has additionally incorporated two other exonic polymorphisms at positions +1192 (exon7) and +1719 (exon 11). These SNPs, identified in the extracellular 3^rd^ and 5^th^ Ig-like domains of the KDR gene receptor, lead respectively to an amino acid changes at residues 297 V>I and 472H>Q and influence VEGF binding efficiency. Indeed, the 3^rd^ domain is critical for ligand fixation while the 5^th^ domain is necessary for VEGF retention on KDR^25^. The genotype and allele distributions of +1192G/A in patients are very close to that found in controls. This substitution did not appear to influence Scz as the difference is statistically insignificant (*p*>0.05). Concerning the +1719A/T polymorphism, T allele and TT genotype (under the three genetic models) was related to the risk of developing this psychosis and more specifically to the paranoid subtype. Previously, our lab results highlighted genetic associations of IFNγ^53^, TNFα/β^54^ and IL-8^55^ particularly for the paranoid amongst the undifferentiated and disorganized clinical forms. In connection with the above data, haplotypic analysis exhibited a higher prevalence of +1192/+1719 GT haplotype in patients when compared to the wild type combination. Similar result with a stronger association is obtained for the paranoid group. Our data are consistent with the functional analysis of these two non-synonymous variants. In fact, lowered mRNA levels generated by the minor +1719T allele are plausibly caused by altering the splicing mechanism due to its proximity (3bp) to the intron/exon boundary. Further, in HEK293 cells, +1719A/T conveyed a 46% augmentation in KDR phosphorylation after VEGF-A165 stimulation (*p*=0.035) while the other substitutions like +1192G/A displayed no effect^56^. Moreover, when we compared the combined effect of the VEGF +936C/T and KDR +1719A/T polymorphisms we noted that the observed predisposing effect may be driven by the VEGF+936T allele. However, among the paranoid subgroup the +936/+1719 TT haplotype carrying two mutated alleles conferred the highest significant difference as compared to the wild type combination.

The attenuated KDR levels in the PFC are inversely associated with positive symptoms of Scz ^49,57^. This receptor is expressed in neurons and blood vessels of the adult human brain^58^. Neurotrophic factors such as BDNF, neuroregulins and EGFs are involved in the pathogenesis of Scz^59,60^ and reduced KDR expression in patients can provoke altered neurotrophic signaling in neuronal cells. Microcirculation anomalies are also linked to Scz^61^. Indeed, analysis performed *post-mortem* revealed ultrastructural malformations, such as thickening of the capillary basement membrane and swelling of pericapillary astrocytic end-foot in the PFC and the visual cortex. As well, imaging studies exhibited larger retinal venules causing microvascular dysfunctions^62^. Thus, the impact of decreased KDR binding affinity in PFC can disseminate to Nacc and cause positive symptoms via glutamatergic neurons^57^. Besides, KDR inhibitors such as *sunitinib* and *sorafenib* generate acute and transient side effects similar to positive symptoms including visual and auditory hallucinations, paranoid delusions and aggressive behavior^63,64,65^. Other reports denoted that elevated VEGF serum levels are negatively correlated with total volume of the frontal lobe in schizophrenic subjects^66^. While stimulation of KDR by VEGF promotes its internalization and degradation^67^, reduced KDR levels in Scz might reflect its constitutive activation^57^.

To date, the link between KDR signaling and susceptibility to Scz remains poorly elucidated. Selective invalidation experiments of, either the receptor or the Src family kinases, have demonstrated the involvement of the VEGF/KDR/Src kinase pathway in the guidance process. During embryonic development, KDR allows guidance of subicular axons in a VEGF-independent manner. Moreover, KDR binds with Plexin D1/Nrp-1 complex and stimulates PI3K/Akt pathway in subicular neurons after fixation of Sema3E^68^. In migrating granule cells, VEGF modulates the association of KDR with NMDA receptor which facilitates calcium entry by the intermediary of Src kinases^69^.

These different data highlight the complexity of KDR signaling that uses distinct pairs of receptors to interact with the nervous system independently of the vascular system.

## Data Availability

No dataset was used to support this study.

## Data Availability

No dataset was used to support this study.

## Disclosure

The authors alone are responsible for the content and writing of the paper.

## Conflicts of Interest

The authors declare that they have no conflicts of interest.

## Acknowledgments

We deeply give thanks to all patients and healthy controls with whose cooperation this study was possible. We also gratefully acknowledge the crew of the Psychiatry Department of Monastir Hospital and the staff of the regional blood transfusion center of the same University Hospital for providing samples and clinical information. All financial support has been provided by the Ministry of Higher Education, Scientific Research and Technology of Tunisia.

## Notes

### Competing Interest Statement

The authors have declared no competing interest.

